# Beneficial Effect of Corticosteroids in Preventing Mortality in Patients Receiving Tocilizumab to Treat Severe COVID-19 Illness

**DOI:** 10.1101/2020.08.31.20182428

**Authors:** Manuel Rubio-Rivas, Mar Ronda, Ariadna Padulles, Francesca Mitjavila, Antoni Riera-Mestre, Carlos García-Forero, Adriana Iriarte, Jose M. Mora, Nuria Padulles, Monica Gonzalez, Xavier Solanich, Merce Gasa, Guillermo Suarez, Joan Sabater, Xose L. Perez-Fernandez, Eugenia Santacana, Elisabet Leiva, Albert Ariza-Sole, Paolo D. Dallaglio, Maria Quero, Antonio Soriano, Alberto Pasqualetto, Maylin Koo, Virginia Esteve, Arnau Antoli, Rafael Moreno, Sergi Yun, Pau Cerda, Mariona Llaberia, Francesc Formiga, Marta Fanlo, Abelardo Montero, David Chivite, Olga Capdevila, Ferran Bolao, Xavier Pinto, Josep Llop, Antoni Sabate, Jordi Guardiola, Josep M. Cruzado, Josep Comin-Colet, Salud Santos, Ramon Jodar, Xavier Corbella

## Abstract

**Introduction:** On the basis of the preliminary report from the RECOVERY trial, the use of dexamethasone or alternative corticosteroids (CS) is currently recommended in severe COVID-19 patients requiring supplemental oxygen. However, last updated recommendations have not taken a position either for or against the use of other immunomodulators such as tocilizumab (TCZ), with or without CS, since results are still limited.

**Methods:** From March 17 to April 7, 2020, a real-world observational retrospective analysis was conducted at our 750-bed university hospital to study the characteristics and risk factors for mortality in patients with severe COVID-19 treated with TCZ +/-CS, in addition to standard of care (SOC). Data were obtained from routine clinical practice, stored in electronic medical records. The main outcome was all-cause in-hospital mortality.

**Results:** A total of 1,092 COVID-19 patients were admitted during the study period. Of them, 186 (17%) were treated with TCZ, of which 129 (87.8%) in combination with CS. Of the total 186, 155 (83.3 %) patients were receiving non-invasive ventilation when TCZ +/-CS was initiated. Mean time from symptoms onset and hospital admission to TCZ use was 12 (± 4.3) and 4.3 days (± 3.4), respectively. Overall, 147 (79%) survived and 39 (21%) died. By multivariate analysis, mortality was associated with older age (HR=1.09, p<0.001), chronic heart failure (HR=4.4, p=0.003), and chronic liver disease (HR=4.69, p=0.004). The use of CS, in combination with TCZ, was the main protective factor against mortality (HR=0.26, p<0.001) in such severe COVID-19 patients receiving TCZ. No serious superinfections were observed after a 30-day follow-up.

**Conclusions:** In severe COVID-19 patients receiving TCZ due to systemic host-immune inflammatory response syndrome, the use of CS in addition to TCZ therapy, showed beneficial effect in preventing in-hospital mortality.

---

As of August 24, 2020, the novel coronavirus (SARS-CoV-2) has infected 23 million people worldwide, and killed more than 800,000 persons [1,2]. The 2019 COVID pandemic was confirmed to have spread to Europe on January 31, 2020 [3]. Since then, there have been more than 400,000 confirmed cases in Spain, with more than 28,000 deaths. As a result, Spain saw one of the most draconian Covid-19 blockades in Europe, but two months after its lift, the country is on the brink of a second wave of coronavirus infections.

Whereas the pandemic continues to spread globally, a worrying 15% of patients will continue to transit into the third and most severe stage of disease [4]. Unlikely to the early clinical stages in which viral replication and local respiratory involvement seem to be the norm, the advanced stage of COVID-19 appears to be triggered by the host-immune response. This third clinical stage presents as severe pulmonary injury and cytokine release syndrome (CRS) with elevation of multi-organ inflammatory markers [4,5].

Accordingly, to treat this advanced stage of COVID-19 illness, the use of immunomodulatory agents such as corticosteroids (CS) or tocilizumab (TCZ), an anti-IL-6 monoclonal antibody that has proven effective in IL-6 mediated diseases, may be justified and has been suggested [6-8]. Currently, on the basis of the preliminary report from the RECOVERY trial, last updated (July 30, 2020) COVID-19 treatment guidelines recommend the use of dexamethasone, or alternative glucocorticoids [9,10]. These immunomodulatory agents are indicated in severe COVID-19 patients requiring supplemental oxygen, being or not mechanically ventilated. Conversely, since studies are still limited, current international recommendations have not taken a position either for or against the use of TCZ in such patients [11].

In this regard, after the first short series of 21 patients reported by Xu et al. [7], the published evidence for the use of TCZ in severe COVID-19 illness has been summarized in two existing systematic reviews and meta-analysis (SRMA). Because the lack of RCTs [12], both SRMA only included observational studies, in which TCZ was given at a single dose of 400-800 mg iv or 162-324 mg sc, renewable once in case of insufficient response to therapy. The first SRMA, published by Lan SH Zhang *et al*. [13], included 7 studies, with no conclusive evidence that TCZ would provide any additional benefit to patients with severe COVID-19. The second, registered in the medRxiv repository by Boregowda *et al*. [14] included 16 studies, concluding that the addition of TCZ to the SOC might reduce the mortality in severe COVID-19.

Nowadays, there is an emerging number of additional observational studies of higher quality from Italy, Spain, France and the US, either recently published or registered in the medRxiv repository [15-24]. These studies mostly assessed the use of TCZ in the subset of severely-ill non-intubated COVID-19 patients and were compared to a control group. Most of these recent studies concluded that TCZ may reduce ICU admissions, mechanical ventilator use, and risk of death. Since most of the studies were performed before the RECOVERY trial, there was no standard protocol in place regarding the use of CS in COVID-19. Thus, the use of CS only depended on the individual decision of those physicians who cared for the included patients. The potential effect of CS, in regimen combination with TCZ, was not specifically evaluated in most of such studies.

This fact prompted us to review our real-world observational data, collected from routine clinical practice, during the first wave of SARS-CoV-2 infections occurred in March-April, 2020, at our hospital setting. Our aim was to compare survivor and non-survivor groups from a large cohort of severe COVID-19 patients treated with TCZ, either alone or in combination with CS, in order to assess the characteristics and associated risk factors for in-hospital mortality in such patient population receiving TCZ.

## METHODS

The first patient with advanced stage of COVID-19 with respiratory failure and systemic host-immune response treated with TCZ at the Bellvitge University Hospital, was on March 17, 2020. From March 17, 2020, to April 7, 2020, during the first 3 weeks of the epidemic, a retrospective cohort study was conducted at our 750-bed tertiary care public institution for adults in Barcelona, Spain. The aim was to describe the characteristics of patients receiving TCZ, with or without concomitant CS, for treating advanced stage of COVID-19, and to assess its effect in preventing all-cause in-hospital mortality. Demographic and clinical data were collected from institutional electronic medical records.

### Inclusion criteria

All patients aged ≥ 18 years admitted at our hospital and given TCZ due to severe COVID-19 pneumonia and systemic hyperinflammation were included consecutively. All patients were diagnosed with COVID-19 by positive polymerase-chain reaction (PCR) of nasal or pharyngeal swabs. All patients underwent a chest X-ray and were reported as COVID-19 lung involvement by an expert radiologist. An electrocardiogram was also performed on all patients on admission and every 48-72 hours during their stay in hospital. Patients admitted to either conventional hospital wards, semi-critical (noninvasive mechanical ventilation) or critical care units (invasive mechanical ventilation) were included.

For TCZ to be used, according to hospital guidelines, patients had to meet a PaO2 (mmHg)/FiO2 (%) x100 <300, or its surrogate SatO2 (%)/FiO2 (%) x100 <315 [25], and at least 2 of the following criteria: ferritin >1000 ng/ml, C-reactive protein (CRP) >100 mg/l, interleukin-6 (IL6) >70ng/l, D-dimer >1000 mcg/l or lactate dehydrogenase (LDH) >400 U/l. An analysis of these inflammatory parameters was performed on the day before administration of the initial dose of TCZ (D0), and subsequently on day 1 (D1), 2 (D2), and 7 (D7) after administration. Lymphocyte count was additionally recorded.

### Outcome

The primary outcome was all-cause in-hospital mortality.

### Variables

As mentioned above, ancillary laboratory tests included PaO2 (mmHg), ferritin, CRP, IL-6, D-dimer, LDH, and lymphocyte count. Myocarditis was defined by clinical criteria, accompanied by sinus tachycardia, elevated troponin T (cut-off ≥14 ng/l) and elevated N-terminal pro-brain natriuretic peptide (NT-proBNP, cut-off ≥300ng/l). Pulmonary thromboembolism (PE) was diagnosed when patients presented with disproportionate dyspnea with D-dimer elevation, and PE was confirmed by CT scan. Low-flow oxygen was defined as nasal cannula 2-3 l or oxygen mask with FiO2 <50%. High-flow oxygen was defined as Monaghan® oxygenation with FiO2 100% and >10l. High-flow oxygen with nasal cannula (HFNC) was defined as nasal cannula oxygenation with FiO2 >50%. For non-invasive ventilation, patients were admitted in semi-critical or intensive care units (ICU). If invasive ventilation with endotracheal intubation was required, patients were admitted at ICU.

### Drug Dosing and Regimens

During the SARS-CoV-2 epidemic in Spain, the use of TCZ in severe COVID-19 patients was regulated, authorized and centrally supplied by the Spanish Ministry of Health, as a single iv infusion at a dose of 400 mg (weight <80kg) or 600 mg (weight >80kg), in order to control TCZ nationwide pharmacological stock. Exceptionally, if there was a partial or incomplete clinical response, with persistence of fever and worsening of analytical parameters, a second or a third infusion of TCZ at 400 −600 mg at 12 hours after the first infusion or a third at 24 hours after the second one, respectively, could be requested from the Spanish Central Government, but authorized in only very few cases per hospital. Regarding the use of CS, there was no standard protocol in place for its use during the study period of the present revision. CS were indicated in combination with TCZ in the majority of cases, depending on individual physicians, using methylprednisolone at doses ranging from 0.5 mg/kg/d to 250mg iv in 3 pulses.

Standard of care (SOC) treatments additionally administered to TCZ and CS were in accordance with available hospital guidelines at the time of the study period (March-April, 2020), and were also evaluated. SOC included hydroxychloroquine (HCQ), azithromycin (AZI), lopinavir/ritonavir (L/r), and remdesivir (R), alone or in combination regimens, given at standard recommended doses [26-29]. Prophylactic anticoagulation therapy with low molecular weight heparin (LMWH) was also recommended. A subset minority of severe COVIS-19 patients received sc interferon beta-1b (IFN) at 0.25mg/48h.

Informed consent was waived because the study was performed outside the context of RCT, and was retrospective in nature. Data were obtained from routine daily practice and anonymized. The confidential information of the studied patients was protected according European and National normative. The study received the ethics approval by the Research Ethics Committee of Bellvitge University Hospital (PR145/20).

### Statistical Analysis

Categorical variables were shown as absolute numbers and percentages. Quantitative variables were shown as mean and standard deviation. Categorical variables were compared using chi-square or Fisher’s test. Quantitative variables were compared using t-test or Mann-Whitney U test depending on whether the variable followed a parametric distribution or not. The Cox proportional-hazards model was performed with mortality as a dependent variable and following a backwards stepwise selection of variables. The survival time used in Cox regression is the time from hospital admission (usually coinciding with the start of the first treatment administered) to the last visit. All variables with significance <0.05 in the univariate study plus age and gender were included in the multivariate study.

## RESULTS

### General data

Between March 17 and April 7, 2020 there were 1,092 patients admitted to our hospital for COVID-19 illness. Of them, 186 (17%) were treated with TCZ at any time of their admission and were included in the study; 129 were male (69.4%), most of old age (64.3 yr. ± 13). At the time of TCZ first infusion, 123 (66.1%) patients were admitted into conventional hospital wards, and 63 (33.9%) into semi-critical/critical care units. Of the total of 186 patients, 147 (79%) survived and 39 (21%) died. Both groups of patients, survivors and non-survivors, were comparable in the baseline. However, patients from the group of non-survivors were older (62.1 years vs. 72.4, p<0.001), more frequently had a previous diagnosis of chronic heart failure (1.4% vs. 12.8%, p=0.005) and/or chronic liver disease (2% vs. 10.3%, p=0.036). Significantly, survivors received CS in combination with TCZ in greater proportion than non-survivors (87.8% vs. 69.2%, P=0.021) (Table 1).

**Table 1.** Demographic, clinical, laboratory and radiographic findings of COVID-19 patients receiving TCZ +/-CS. Comparison between survivor and non-survivor groups.

|  | **All patients**<br>**N=186** | **Survivors**<br>**N=147** | **Non-survivors**<br>**N=39** | **p-value** |
| --- | --- | --- | --- | --- |
| **Age, years mean (SD)** | 64.3 (13) | 62.1 (12.7) | 72.4 (10) | **<0.001** |
| **Gender (males), n (%)** | 129 (69.4) | 103 (70.1) | 26 (66.7) | 0.682 |
| **Body mass index, mean (SD)** | 29.3 (4.1) | 29.6 (4.2) | 28.4 (3.6) | 0.105 |
| **Smoking behavior, n (%)** |  |  |  |  |
| Never smoker | 140 (75.3) | 108 (73.4) | 32 (82) | 0.532 |
| Current smoker | 8 (4.3) | 7 (4.8) | 1 (2.6) |  |
| Former smoker | 38 (20.4) | 32 (21.8) | 6 (15.4) |  |
| **Comorbidity n (%)** |  |  |  |  |
| Arterial hypertension | 92 (49.5) | 68 (46.3) | 24 (61.5) | 0.090 |
| Diabetes mellitus | 45 (24.2) | 32 (21.8) | 13 (33.3) | 0.134 |
| Hyperlipidemia | 84 (45.2) | 62 (42.2) | 22 (56.4) | 0.112 |
| COPD* | 8 (4.3) | 7 (4.8) | 1 (2.6) | 1.000 |
| Asthma | 7 (3.8) | 6 (4.1) | 1 (2.6) | 1.000 |
| Chronic heart failure | 7 (3.8) | 2 (1.4) | 5 (12.8) | **0.005** |
| Ischemic cardiopathy | 11 (5.9) | 7 (4.8) | 4 (10.3) | 0.246 |
| Atrial fibrillation | 6 (3.2) | 3 (2) | 3 (7.7) | 0.108 |
| Chronic kidney disease | 16 (8.6) | 10 (6.8) | 6 (15.4) | 0.089 |
| Chronic liver disease | 7 (3.8) | 3 (2) | 4 (10.3) | **0.036** |
| Cancer | 18 (9.7) | 12 (8.2) | 6 (15.4) | 0.175 |
| Autoimmune disorder | 14 (7.5) | 10 (6.8) | 4 (10.3) | 0.497 |
| **Symptoms n (%)** |  |  |  |  |
| Fever >38°C | 174 (93.5) | 140 (95.2) | 34 (87.2) | 0.069 |
| Cough | 144 (77.4) | 110 (74.8) | 34 (87.2) | 0.101 |
| Dyspnea | 117 (62.9) | 93 (63.3) | 24 (61.5) | 0.843 |
| Anosmia | 18 (9.7) | 15 (10.2) | 3 (7.7) | 0.769 |
| Ageusia | 21 (11.3) | 17 (11.6) | 4 (10.3) | 1.000 |
| Sore throat | 14 (7.5) | 12 (8.2) | 2 (5.1) | 0.738 |
| Diarrhea | 63 (33.9) | 53 (36.1) | 10 (25.6) | 0.222 |
| Arthromyalgia | 56 (30.1) | 41 (27.9) | 15 (38.5) | 0.201 |
| **Myocarditis/MI<sup>†</sup>, n (%)** | 1 (0.5) | 1 (0.7) | 0 | 1.000 |
| **Pulmonary thromboembolism, n (%)** | 9 (4.8) | 6 (4.1) | 3 (7.7) | 0.399 |
| **PaO<sub>2</sub>/FiO<sub>2</sub> D<sub>0</sub>, mean (SD)/N** | 188.4 (104.1)/137 | 194 (106)/107 | 169 (96)/30 | 0.237 |
| **SatO<sub>2</sub>/FiO<sub>2</sub> D<sub>0</sub>, mean (SD)/N** | 175.7 (80.1)/129 | 182.6 (84.8)/101 | 151 (54.5)/28 | **0.021** |
| **PaO<sub>2</sub>/FiO<sub>2</sub> <300 or SatO<sub>2</sub> <315 D<sub>0</sub>, n (%) /N** | 155 (83.3)/186 | 119 (81)/147 | 36 (92.3)/39 | 0.144 |
| **Maximum Oxygen/ventilation required, n (%)** |  |  |  |  |
| Low-flow oxygen | 5 (2.7) | 5 (3.4) | 0 | 0.200 |
| High-flow oxygen (Monaghan®) | 9 (4.8) | 8 (5.4) | 1 (2.6) |  |
| High-flow oxygen (HFNC<sup>‡</sup>) | 7 (3.8) | 7 (4.8) | 0 |  |
| Noninvasive ventilation | 155 (83.3) | 121 (82.3) | 34 (87.2) |  |
| Invasive ventilation | 10 (5.4) | 6 (4.1) | 4 (10.3) |  |

\* COPD: chronic obstructive pulmonary disease. <sup>†</sup>MI: myocardial infarction <sup>‡</sup>HFNC: high flow nasal cannula

### Ancillary tests

Inflammatory parameters and lymphocyte count documented the day before TCZ first administration (D0), with or without CS, and its progression during the following 7 days (D1, D2, and D7) are shown in Table 2. Although daily evolution of laboratory data was registered in a limited number of included patients, survivors showed much lower levels of abnormal inflammatory parameters and higher lymphocyte count the day before TCZ first infusion when compared to non-survivors. Furthermore, non-survivors showed a higher rapid increase of IL-6 two days after TCZ infusion (D2), as well as an evolution to worse of LDH plasmatic levels. However, none of these differences were found to be statistically significant for predicting in-hospital mortality in this study.

**Table 2.** Laboratory tests: Inflammatory parameters and lymphocyte count. Comparison between survivor and non-survivor groups.

|  | D0 | D1 | D2 | // | D7 |
| --- | --- | --- | --- | --- | --- |
| **All patients, mean (SD)/N**<br>**(N=186)** |  |  |  |  |  |
| Ferritin ng/ml | 1,927(1,245)/89 | 1,789(1,213)/76 | 1,853(1,478)/75 |  | 1,211(624)/77 |
| CRP mg/l | 188(142)/148 | 133(106)/116 | 66(53)/112 |  | 21(50)/108 |
| IL6 ng/l, | 191(450)/32 | 700(1,243)/28 | 1,184(1,817)/19 |  | 665(711)/14 |
| D-dimer mcg/l | 2,258(6,026)/131 | 3,615(7,701)/108 | 3,717(7,391)/107 | // | 2,253(2,268)/106 |
| LDH U/l | 459(154)/136 | 469(189)/109 | 459(184)/108 |  | 455(236)/108 |
| Lymphocytes x10<sup>6</sup>/l | 900(1,509)/184 | 720(375)/113 | 850(873)/116 |  | 1,300(1,085)/105 |
| **Survivors, mean (SD)/N**<br>**(N=147)** |  |  |  |  |  |
| Ferritin ng/ml | 1,911(1,250)/74 | 1,764(1,195)/65 | 1,854(1,485)/67 |  | 1,142(569)/70 |
| CRP mg/l | 175(110)/121 | 127(105)/94 | 62(52)/91 |  | 21(53)/98 |
| IL6 ng/l, | 188(507)/25 | 657(1,328)/24 | 688(1,112)/17 | // | 495(445)/12 |
| D-dimer mcg/l | 1,682(3,704)/109 | 2,465(5,237)/87 | 3,034(6,114)/87 |  | 2,042(2,017)/96 |
| LDH U/l | 442(141)/114 | 440(158)/88 | 430(167)/88 |  | 421(211)/97 |
| Lymphocytes x10<sup>6</sup>/l | 820(396)/146 | 740(380)/92 | 910(939)/95 |  | 1,370(1,119)/94 |
| **Non-survivors, mean (SD)/N**<br>**(N=39)** |  |  |  |  |  |
| Ferritin ng/ml | 2,006(1,258)/15 | 1,936(1,370)/11 | 1,837(1,518)/8 |  | 1,898(782)/7 |
| CRP mg/l | 246(232)/27 | 161(110)/22 | 83(55)/21 |  | 12(15)/10 |
| IL6 ng/l, | 200(132)/7 | 956(533)/4 | 5,395(24)/2 | // | 1,683(1,403)/2 |
| D-dimer mcg/l | 5,113(11,999)/22 | 8,378(13,031)/21 | 6,691(11,140)/20 |  | 4,280(3,475)/10 |
| LDH U/l | 551(187)/22 | 589(257)/21 | 587(203)/20 |  | 751(243)/11 |
| Lymphocytes x10<sup>6</sup>/l | 723(431)/38 | 548(352)/21 | 548(352)/21 |  | 715(416)/11 |
| *CRP: C-reactive protein. <sup>†</sup>IL6: interleukin-6. <sup>‡</sup>LDH: lactate dehydrogenase. |  |  |  |  |  |

### Drugs combinations and doses

All drugs in this study were given in accordance with the standard hospital protocol during the study period, based on recommended international guidelines available at that time. The number of patients receiving each of the drugs administered and the duration of such treatments is detailed in Table 3.

**Table 3.**
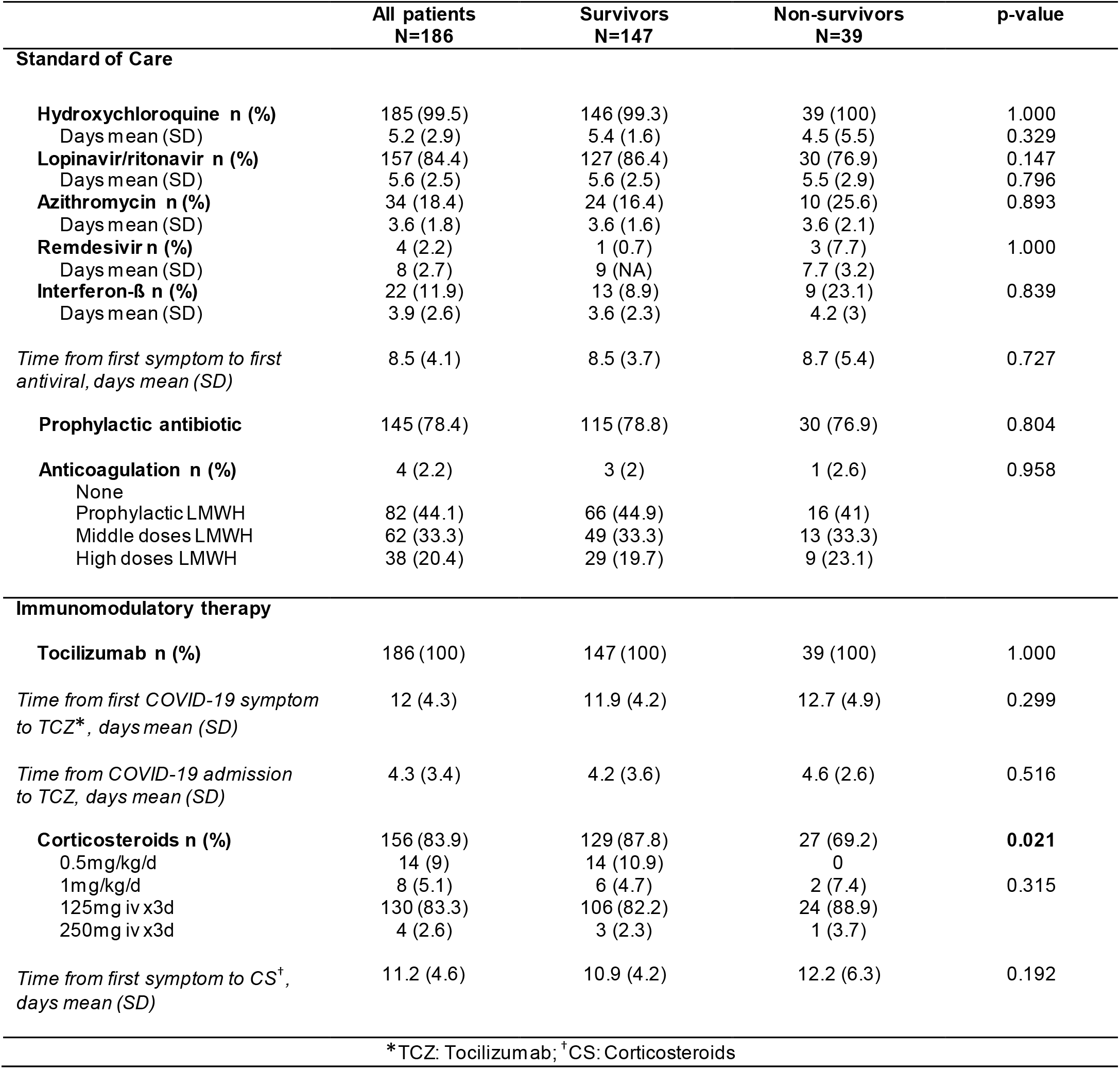
Drugs, doses and duration. Comparison between survivor and non-survivor groups.

The most common drugs used as SOC were HCQ (99.5%) and L/r (84.4%). The usual sequential regimen was an initial treatment with a combination of HCQ plus L/r for 5 days given on day +8.5 (± 4.1) from the beginning of COVID-19 symptoms. The number of patients treated with prophylactic antibiotic (generally iv amoxicillin-clavulanate, piperacillin-tazobactam or ceftriaxone) and anticoagulation therapy are also indicated in Table 3.

In those patients at risk to transit to a more severe stage, this initial regimen was subsequently followed by CS (iv pulses of 125 mg x 3 days) given on day +11.1 (± 4.6) and followed by or overlapped with TCZ (single dose) given on day +12 (± 4.3). In this regard, CS were also administered in combination with TCZ in 156 (83.9%) of the total 186 included patients. There were no differences between survivor and non-survivor groups on the doses of CS received. Nor was there any difference in the sequential order of infusion between the TCZ and CS. Regarding TCZ administration, most patients (92.5%) received a single infusion of TCZ, 13 (7%) a second infusion 12h after the first one, and in only 1 case (0.5%) a third infusion was administered 24h after the second infusion. In 14 of the total 186 (7.5%) patients, despite presenting a PaO2/FiO2 greater than 300, TCZ prescription was indicated according to the serious alteration of inflammatory parameters and lymphocyte count. No noticeable adverse events related to TCZ +/-CS were shown throughout the study period and within the first 30 days of follow-up.

Finally, no differences between both survivor and non-survivor groups were shown according to both the indication of anticoagulation and the different doses received, showing no higher mortality protection in the univariate study.

### Risk factors of mortality

When survivors and non-survivors were compared in COVID-19 patients receiving TCZ +/-CS due to severe lung injury and systemic host-immune response, the multivariate study showed statistical significance for older age (HR=1.09, CI 95% 1.05-1.13, p<0.001), previous chronic heart failure (HR=4.4, CI 95% 1,68-11.63, p=0.003), and previous chronic liver disease (HR=4.69, CI 95% 1.62-13.59, p=0.004) as risk factors for all-cause in-hospital mortality. In contrast, the use of CS in combination with TCZ therapy showed a beneficial effect in preventing mortality in such subset of COVID-19 patients receiving TCZ (HR=0.26, CI 95% 0.13-0.56, p<0.001) (Table 4).

**Table 4.** Risk factors associated with all-cause in-hospital mortality in COVID-19 patients receiving TCZ +/-CS. Cox regression analysis.

|  | Univariate analysis |  | Multivariate analysis |  |
| --- | --- | --- | --- | --- |
|  | HR (95%CI) | p-value | HR (95%CI) | p-value |
| **Age/year** | **1.07 (1.04-1.11)** | **<0.001** | **1.09 (1.05-1.13)** | **<0.001** |
| **Gender (male)** | 0.77 (0.39-1.51) | 0.445 | — | — |
| **BMI\*** | 0.95 (0.87-1.03) | 0.205 |  |  |
| **Smoking behavior** |  |  |  |  |
| Never smoker (ref.) | 1 | - |  |  |
| Current smoker | 0.42 (0.06-3.09) | 0.395 |  |  |
| Former smoker | 0.48 (0.19-1.23) | 0.127 |  |  |
| **Comorbidity** |  |  |  |  |
| Arterial hypertension | 1.59 (0.83-3.04) | 0.166 |  |  |
| Diabetes mellitus | 1.36 (0.68-2.69) | 0.384 |  |  |
| Hyperlipidemia | 1.41 (0.75-2.68) | 0.290 |  |  |
| COPD<sup>†</sup> | 0.43 (0.06-3.13) | 0.404 |  |  |
| Asthma | 0.47 (0.06-3.42) | 0.454 |  |  |
| **Chronic Heart failure** | **5.86 (2.26-15.23)** | **<0.001** | **4.42 (1.68-11.63)** | **0.003** |
| Ischemic cardiopathy | 1.81 (0.64-5.10) | 0.264 |  |  |
| Atrial fibrillation | **4.13 (1.26-13.58)** | **0.020** | — | — |
| Chronic kidney disease | 1.79 (0.70-4.60) | 0.224 |  |  |
| **Chronic liver disease** | **3.12 (1.11-8.81)** | **0.032** | **4.69 (1.62-13.59)** | **0.004** |
| **Cancer** | **2.56 (1.06-6.17)** | **0.036** | — | — |
| Autoimmune disorder | 1.23 (0.44-3.47) | 0.696 |  |  |
| **Lab test D0** |  |  |  |  |
| Ferritin >1500 ng/ml | 0.88 (0.32-2.44) | 0.810 |  |  |
| CRP<sup>‡</sup> >150 mg/l | 2.21 (0.96-5.06) | 0.061 |  |  |
| IL6<sup>§</sup> >100 ng/l, | 6.53 (0.78-54.31) | 0.083 |  |  |
| D-dimer >1500 mcg/l | 1.34 (0.57-3.17) | 0.500 |  |  |
| LDH<sup>¶</sup> >400 U/l | 3.27 (0.96-11.10) | 0.057 |  |  |
| Lymphocytes <600 x10<sup>6</sup>/l | 1.47 (0.77-2.82) | 0.242 |  |  |
| **SatO2/FiO2** | 0.99 (0.99-1) | 0.661 |  |  |
| **PaO2/FiO2 <300** | 1.38 (0.42-4.57) | 0.597 |  |  |
| **Pulmonary thromboembolism** | 1.18 (0.36-3.86) | 0.788 |  |  |
| **Anticoagulation** |  |  |  |  |
| None (ref.) | 1 | - |  |  |
| Prophylactic LMWH<sup>#</sup> | 0.93 (0.12-7.05) | 0.947 |  |  |
| Middle doses LMWH | 1.05 (0.14-8.01) | 0.965 |  |  |
| High doses LMWH | 1.11 (0.14-8.89) | 0.922 |  |  |
| **Time from first symptom to TCZ** | 0.99 (0.95-1.03) | 0.532 |  |  |
| **Number of infusions of TCZ (1 vs. 2-3)** | 1.64 (0.68-3.96) | 0.267 |  |  |
| **Use of CS\*\* in combination with TCZ** | **0.33 (0.16-0.68)** | **0.002** | **0.26 (0.13-0.56)** | **<0.001** |

\*BMI: body mass index. <sup>†</sup>COPD: chronic obstructive pulmonary disease. <sup>‡</sup>CRP: C-reactive protein.
<sup>§</sup>IL6: interleukin-6. <sup>¶</sup>LDH: lactate dehydrogenase. <sup>#</sup>LMWH: low-molecular-weight heparin. <sup>||</sup>TCZ: tocilizumab, \*\*CS: corticosteroids

## DISCUSSION

While a majority of COVID-19 patients show a low-mild disease with an overall case mortality rate of 2-3%, a subset of 15% of patients presented with lung involvement of moderate severity requiring hospital admission, and 5% with severe respiratory failure and systemic host-immune inflammatory response, resulting in fatality in half of such cases [1-3]. These findings are consistent with the three stages of increasing severity proposed by Siddiqu and Mehra: stage I (incubation and early infection), stage II (pulmonary involvement), and stage III (systemic hyperinflammation and multi-organ dysfunction) which correspond with distinct clinical findings and outcomes [4].

In this regard, only remdesivir [28] and dexamethasone [9] have demonstrated evidence-based efficacy on randomized, controlled clinical trials (RCTs) in severe COVID-19 illness to date [10]. These results are in accordance with the above-mentioned natural 3-stage evolution of COVID-19 disease. In this regard, SARS-CoV-2 viral incubation and early establishment of the disease appears to be the predominant component in the first week, viral replication and transition into moderate acute lung involvement the most important in the second one, and severe lung injury and systemic host-immune inflammatory response from the third [4]. Therefore, it seems reasonable to propose a sequential combined therapy with effective antiviral drugs as first step and, thereafter, immunomodulatory treatment in those COVID-19 patients progressing into the advanced stage with severe pulmonary failure and systemic hyperinflammatory syndrome.

While the use of dexamethasone, or alternative CS, is currently recommended as immunomodulatory agents for the treatment of severe COVID-19 patients requiring supplemental oxygen [10], updated recommendations have not taken a position either for or against the use of other immunomodulators such as tocilizumab (TCZ), alone or in combination with CS. The use of TCZ in such COVID-19 patients has been justified on the basis of observational reports assessing the effect of TCZ to treat the cytokine release syndrome (CRS) described to occur in severe COVID-19 illness. However, data from RCT supporting its use have not been reported yet [4-8, 13,14].

In this regard, recent real-world observational studies of high quality from Italy, Spain, France and the US, indicated that TCZ might reduce ICU admissions, mechanical ventilator use, and risk of death, when compared to control group only treated with SOC [15-24]. Moreover, Moreno-García et al. [16] conclude that the beneficial effect of TCZ might be especially shown in the subset of severely-ill non-intubated COVID-19 patients at early stage of the systemic hyperinflammatory response. In this respect, the SRMA by Boregowda et al. [14] found that there was no statistical difference in mortality between TCZ and SOC when steroids are used in the treatment of severe COVID-19 (n=12; pooled OR 0.76 [95% CI 0.47, 1.23; p=0.27). However, when steroid was not used, TCZ group had significantly low mortality when compared to the SOC group (n=4; pooled OR 0.24 [ 95% CI; 0.10-0.54] p<0.01). Interestingly, in a vast majority of these studies assessing TCZ in COVID-19, CS were also given in addition to TCZ, ranging 18-71% of included cases; however, the potential role in combined regimen with TCZ was little evaluated.

These results prompted us to perform the present observational investigation, with the aim to identify those factors determining prognosis in a large real-world cohort of 186 patients receiving TCZ, with and without CS, due to severe COVID-19 illness, hospitalized during our first wave of infections. Our findings showed that, in such severe patients receiving TCZ, in-hospital mortality was associated with older age, and prior chronic heart failure or liver disease. These mentioned factors were in accordance with those predicting poor prognostic already found and previously reported in general population with COVID-19 illness.

Of interest, the present study showed that in a vast majority (83.9%) of our severe COVID-19 patients treated with TCZ, TCZ was also given in combination regimen with CS. Remarkably, multivariate analysis identified the addition of CS to TCZ to be protective for in-hospital mortality in our series. In this respect, our results are in line of those reported in the SRMA by Boregowda et al., which concluded that when steroids are used in the SOC, the absence of a significant difference in mortality between those COVID-19 patients receiving TCZ and those who not, suggests a beneficial effect of the anti-inflammatory properties of steroids in the treatment of severe COVID-19.

In the present study, TCZ and CS were both given as second-step therapy after SOC, when the treating physicians considered that patients were probably at higher risk to transit into the advanced stage of COVID-19. Among the 153 included patients receiving TCZ and CS in combination, CS were most commonly given as iv pulses of methylprednisolone at 125 mg x 3 days at day +11, immediately followed by or overlapped with iv TCZ at day +12, at 8 mg/Kg (single dose). Only a minority of our patients (7.5%) received a second or third TCZ infusion, not appearing to be related to lower mortality. Lastly, in contrast with some previous studies in which adverse events could not be evaluated due to short follow-up, we assessed treatment safety during a 30-day follow-up after TCZ +/-CS administration, with no obvious serious adverse events or superinfections.

Finally, although survivors showed much better evolution of abnormal inflammatory parameters and lymphopenia than non-survivors, none of them were found to be statistically significant for predicting prognosis within clinical practice. However, in the limited subset of included patients in whom laboratory data were available throughout the first days after TCZ prescription, it is noteworthy that non-survivors showed a worrying higher rapid increase of IL-6 two days after TCZ infusion, as well as worse evolution of LDH levels. The clinical relevance of these results might be assessed in future studies in order to provide better help to physicians treating patients at severe stage of COVID-19 disease.

### Limitations

The results of the present study show several limitations that must be noted. First, the study is retrospective in nature using real-world observational data, outside the context of RCT. Second, we aimed to evaluate predictors of mortality in the subset of patients receiving TCZ, with or without CS, but not to assess the efficacy of TCZ in general population with COVID-19 illness, comparing treated and untreated groups. Perhaps for this reason, the risk factors showed in our study may differ slightly from those previously recognized in other observational studies [1,30]. Third, due to the emergency situation, small variations to the standard clinical management of COVID-19 patients may not be totally ruled out as result from hospital reorganization, involving a wide spectrum of departments and physician specialists. Fourth, the number of COVID-19 patients presenting with PE and/or myocarditis may have been underestimated, since symptoms are difficult to ascertain in severe COVID-19 patients and the d-dimer and troponins are almost universally elevated. Finally, daily evolution of inflammatory parameters throughout the first days after TCZ administration was registered in a limited number of included patients, which does not allow firm conclusions to be drawn.

### Conclusions

As most physicians worldwide, whereas the pandemic continues to spread globally and our country is waiting for a second wave, we still tackle severe COVID-19 utilizing a variety of immunomodulatory drugs on the only basis of available reports providing valuable observational data, to come up with efficacious solutions quickly. In our large series of 186 consecutive COVID-19 patients receiving TCZ for severe systemic host-immune inflammatory response syndrome, multivariate analysis identified those patients of older age, with a previous diagnosis of chronic heart failure or liver disease to be at higher risk for in-hospital mortality. Conversely, the addition of CS to TCZ therapy was shown to be protective in preventing in-hospital mortality. Unfortunately, on the basis of the present study, it cannot be stated when would be the best time, not too early nor too late, the best regimen and appropriated doses, to use immunomodulatory agents during the clinical evolution of the COVID-19 illness. In this respect, close monitoring of inflammatory markers, before and after treatment, may lead clinicians and researchers conducting RCTs, to select the right patients in whom immunomodulators such as TCZ and CS may be beneficial.

## Data Availability

The datasets generated during and/or analyzed during the current study are available from the corresponding author on reasonable request.

## Acknowledgments

We are indebted to all physicians, nurses and rest of healthcare personnel of Bellvitge University Hospital, University of Barcelona, who have been involved in the care of patients with COVID-19. We dedicate this work to the memory of those patients at the severe stage who did not survive to the COVID-19 illness. No funding or sponsorship was received for this study or publication of this article. The authors declare that they have no conflict of interest. The datasets generated during and/or analyzed during the current study are available from the corresponding author on reasonable request.

